# VR implementation in mental healthcare: A marathon, not a sprint – A qualitative longitudinal evaluation of a VR training program

**DOI:** 10.1101/2025.07.02.25330712

**Authors:** Marileen M.T.E. Kouijzer, Laura A.M. Koenis, David Huizinga, Saskia M. Kelders, Yvonne H.A. Bouman, Hanneke Kip

**Affiliations:** Department of Technology, Human and Institutional Behavior, Centre for eHealth and Wellbeing Research, University of Twente, Enschede, The Netherlands; Department of Research, Transfore, Deventer, The Netherlands; Dimence Mental Health Care Centre, Deventer, The Netherlands; Opentia Research Unit, North-West University, Vanderbijlpark, South Africa

**Keywords:** *Virtual Reality*, *VR*, *Implementation*, *Mental Healthcare*, *TDF*, *COM-B*, *Implementation Strategy*

## Abstract

Despite the potential of Virtual Reality (VR) in mental healthcare, its implementation in practice remains limited. One contributing factor is the lack of research into effective implementation strategies for VR. This longitudinal study examines therapists’ experiences with a VR training program designed as an implementation strategy for VR and explores key factors influencing VR implementation over time.

This longitudinal qualitative evaluation involved eleven therapists from a Dutch mental healthcare organization. The therapists participated in a six-session VR training program and completed semi-structured interviews at three time points: pre-training, post-training, and three months after the training concluded. Data were deductively analyzed using the Capability, Opportunity, Motivation-Behavior model and Theoretical Domains Framework to capture changes in capability, motivation, and opportunity.

The study showed how VR training influenced the implementation process of VR in mental healthcare. Therapists’ self-reported knowledge and skills improved during training, emphasizing the importance of integrating both technical and therapeutic aspects. However, despite improvements in capability, actual VR usage remained limited. Post-training barriers included workflow incompatibility, hierarchical dynamics, and lack of organizational vision and sustained support.

This longitudinal study provided insights into how implementation determinants shifted over time and were influenced by training. While initial training is crucial for developing skills, training alone is insufficient for successful implementation. Organizations must foster a supportive environment, including a clear vision, adequate resources, and an ongoing supportive attitude. The findings highlight the importance of ongoing evaluation and adaptation of implementation strategies over time to ensure sustainable integration of VR into mental healthcare.

**Author Summary:** Virtual reality (VR) offers exciting opportunities to enhance mental healthcare, for example by helping patients practice challenging situations in a safe, controlled environment. But despite its potential, VR is still not widely used in daily clinical practice. In this study, we followed therapists who took part in a VR training program, and interviewed them before, after, and three months after the training. This gave us a unique look at how their experiences and needs evolved over time.

We found that the training helped therapists feel more capable and motivated to use VR. Yet, in practice, many still faced barriers—like technical challenges, lack of time, and minimal organizational support. What stood out is that these barriers often only became clear after the training, once therapists tried to apply VR in real-life settings.

Our study shows that implementing new technologies is not a one-time effort, but a process that unfolds in phases. The factors that influence success shift over time, which means organizations need to keep checking in: what do therapists need now, and how can we support that? By tracking the process over time, our study helps to better understand what sustainable implementation really takes.

## Introduction

### The Use and Implementation of Virtual Reality in Mental Healthcare

Virtual Reality (VR) has emerged as an innovative technology to enhance treatment in mental healthcare. VR offers unique possibilities to improve the treatment of a broad range of mental disorders. It can e.g., be used to improve exposure or skill training, increase the impact of therapy on treatment outcomes, be more cost-effective, and be available to a larger group of patients [1, 2]. VR uses computer-generated interactive environments to imitate real-world situations. This creates the possibility of practicing challenging situations in a safe and controlled environment that can be tailored to the needs or conditions of patients [2]. The potential applications of VR technology in treating anxiety disorders, psychotic disorders, and aggression regulation disorders highlight its promise in enhancing treatment [3–7]. However, despite this potential, the actual usage of VR in clinical settings remains limited [8–11].

The implementation of VR in mental healthcare faces various challenges. Barriers that are often mentioned are the lack of knowledge and skills of therapists regarding the technical aspects of VR use and its integration into treatment [12–18]. One explanation could be that using VR in treatment demands different competencies from therapists than traditional, in-person treatment [19]. For example, therapists not only need sufficient technical skills to use VR, but they also require different therapeutic and content-related skills, such as conducting a role-play in VR and integrating this new technology into their daily workflows [19]. To put these skills into practice, a significant behavior change is asked of these therapists. A qualitative study showed that despite their overall positive attitude towards VR and their intention to integrate it into their practices, they fall short in executing the necessary behaviors for successful integration [19]. This highlights an intention-behavior gap in implementation-related behavior, indicating that despite expressing the intent to change their behavior, therapists may not follow through with concrete actions [20].

One implementation strategy that can address these barriers and possibly bridge the intention-behavior gap is an extensive training program for therapists [19]. In healthcare services, training to integrate new treatment or technology in practice is often limited, creating a barrier to implementation [21]. When training does occur, it typically focuses on increasing the technical skills of practitioners to employ the technology in practice [22, 23]. However, training is often not focused on therapeutic aspects of treatment, such as how to integrate the technology in treatment sessions, how to adapt the technology to patient needs, or how to combine the technology with therapeutic techniques like exposure therapy. The introduction of VR, furthermore, seems to demand a shift in this approach, as it is not merely a technology. VR use fundamentally transforms how treatment is conceptualized and delivered; it requires therapists to adopt new roles, integrate virtual environments into treatment, and rethink traditional treatment structures [2, 24]. These findings suggest that training should go beyond technical skills and include a nuanced understanding of how VR alters the treatment approach and what behavior change may be needed from the involved therapists.

### A Systematic Approach Towards Implementation

Implementation should not be an afterthought but rather be considered from the start of technology design, ensuring alignment with clinical needs and workflows [25]. To approach implementation as a systematic process, tailored implementation strategies, such as a VR training program, should be developed to tackle the barriers effectively [26]. In addition, it is important not only to identify useful implementation strategies and assess their effectiveness but also to understand why some strategies succeed while others fail [26]. It is important to consider determinants of behavior change in implementation. These determinants refer to the underlying factors that influence whether a person adopts or maintains a new behavior, such as their knowledge, beliefs, and external factors like social support [27]. By gaining insight into these factors, we can better tailor strategies to address the specific challenges therapists face. Ensuring they are aligned with the needs of the individuals involved and improving the chances of successful implementation [26, 28].

This is where systematic mapping of determinants of implementation becomes important. To evaluate the implementation of VR in mental healthcare over time, it is recommended to map determinants that influence the implementation process, including therapist behavior, organizational factors, and other contextual elements [29]. The Capability, Opportunity, Motivation-Behavior model (COM-B), which emphasizes capability, opportunity, and motivation as key aspects of behavior change, provides a structured approach to understanding what influences therapists’ implementation of VR technology [30, 31]. This model is often complemented by the Theoretical Domains Framework (TDF), which then breaks these broad categories down into specific determinants, such as knowledge, skills, beliefs in consequences, and social influences [28, 31]. Together, these models can provide both a high-level framework and a deeper understanding of the determinants that impact the implementation of VR [32]. Fig 1 illustrates the integration of both frameworks.

**Fig 1.**
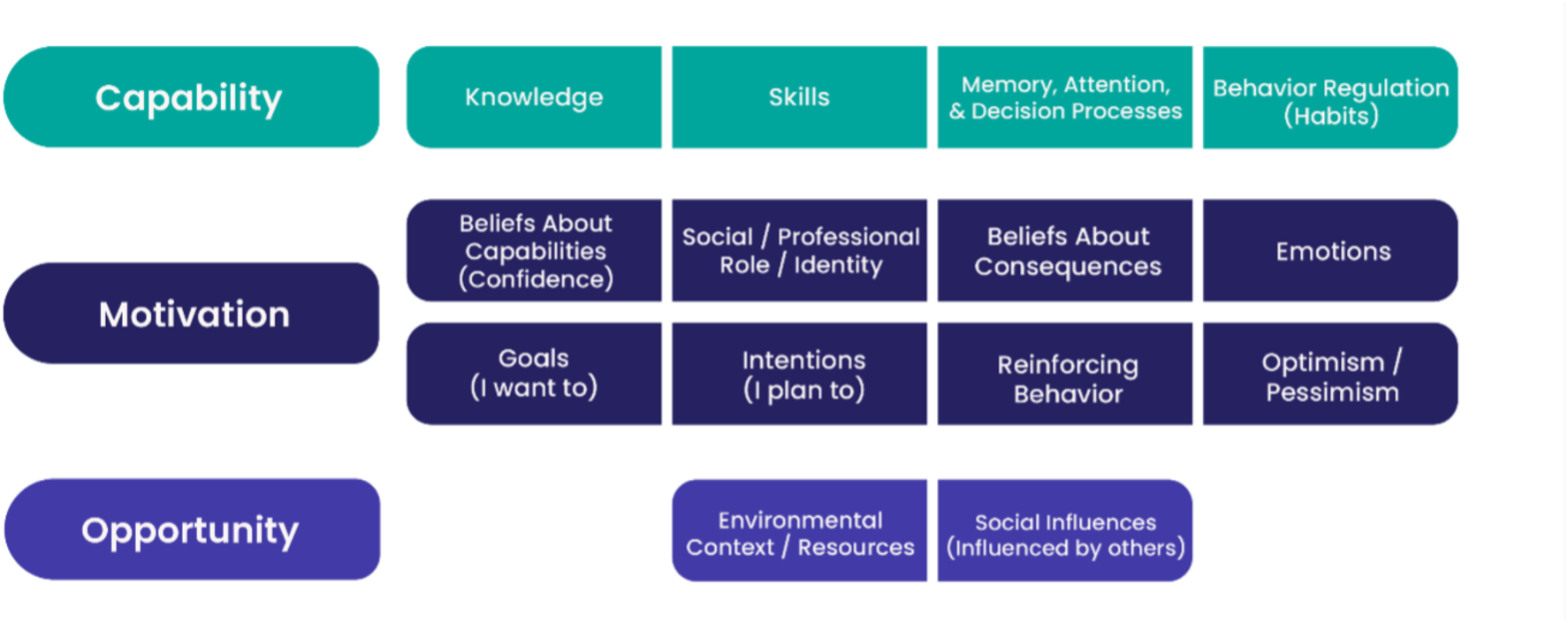
COM-B model and TDF integration based on Michie, van Stralen & West (2011). Image developed by The Center for Implementation ©2023 | V2024.01.

The 14 domains of the TDF framework are split up into the three categories of the COM-B model. The capability category includes TDF domains related to cognitive and practical abilities required by therapists to use VR. These are essential for understanding how therapists can effectively integrate VR technology into their treatment practice. The motivation category includes TDF domains that reflect cognitive and emotional factors influencing motivation to engage in new behavior, such as using VR. The opportunity category includes the TDF domains that capture external factors that can either facilitate or hinder the implementation of VR in practice [32]. The COM-B and TDF complement each other in identifying implementation determinants to change behavior. Their combined use has been effective in various healthcare settings by systematically mapping influencing factors and tailoring implementation strategies accordingly [32, 33].

These models are often used to assess the current situation by identifying barriers and facilitators. However, there is also potential to use them to evaluate implementation strategies. Both frameworks can help assess what changes occur as a result of a strategy, offering insight into which factors have been successfully addressed and which need further attention. The implementation strategy could be adapted accordingly. By mapping these factors in a longitudinal study design, a deeper understanding can be gained of how specific determinants contribute to the implementation of VR and how they change over time in response to the implementation strategies used [34]. Such insights could support the development of targeted, time-sensitive implementation strategies to enhance the effectiveness and sustainability of VR in mental healthcare practice.

### Evaluating the Impact of VR Training on Implementation in Mental Healthcare

The present study seeks to explore how an implementation strategy – a VR training program – impacts therapists’ VR use and their implementation behavior. This longitudinal qualitative evaluation aims to capture how this behavior evolves as therapists gain more experience with VR through the training program. The research questions guiding this study are:

1. *How does a VR training program influence the determinants of VR implementation in mental healthcare, as defined by the TDF and COM-B model, from the perspective of therapists?*
2. *How do these determinants evolve over time, specifically between pre-training, post-training, and follow-up, as therapists gain experience with VR through participation in the VR training program?*

## Methods

### Study Design and Setting

This study was divided into five phases (Fig 2). Three semi-structured interview rounds were conducted with eleven therapists who participated in the VR training program. The five phases consist of; 1) the first interview that took place pre-training and focused on evaluating the needs and expectations of participants; 2) the VR training program that was followed for six sessions over three months; 3) the second interview with therapists that took place directly post-training and elaborated on the experiences of the participants with the training program; 4) the therapists put VR into practice for about three months; and, 5) the third follow-up interview was conducted three months after the VR training program had been concluded. This final interview explored the usage of VR technology in practice and the impact of the VR training program on the implementation of VR technology.

**Fig 2.**
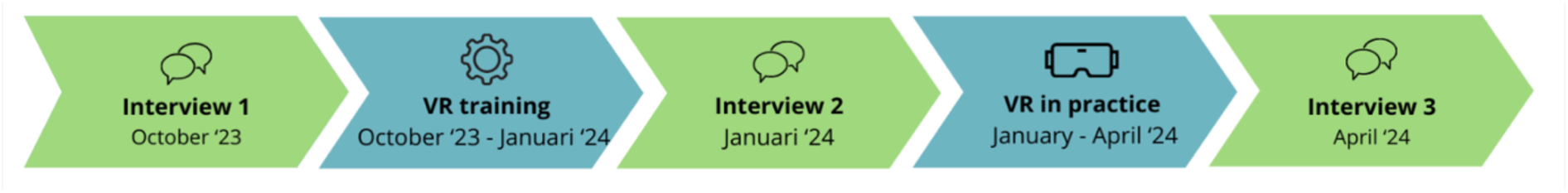
Timeline of the current longitudinal qualitative evaluation study.

This study took place within a mental healthcare organization located in the East of the Netherlands. The organization specializes in providing a wide range of mental healthcare services, catering to both in- and outpatients. This study has been approved by the Ethics Committee of the BMS faculty of the University of Twente (request number 231073). The manuscript follows the Standards for Reporting Implementation Studies (StaRI) guidelines to ensure comprehensive and transparent reporting of implementation research [35].

### VR Technology

In the VR training program, the interactive VR system ‘Social Worlds’ of the Dutch company CleVR was used. This VR software makes it possible to bridge the gap between the real world and the treatment room by exposing patients to a variety of realistic virtual environments and letting them interact with a broad range of virtual characters. The patient can navigate multiple environments, including a supermarket, a shopping street, or a home environment. Furthermore, the patient can engage in role-play with virtual characters, in which these characters are embodied by the therapist using a voice-morphing microphone. The therapist has control over the character’s voice, movements, facial expressions, and body language through a user dashboard. This dashboard allows for dynamic alterations to the virtual surroundings, such as increasing the number of pedestrians on a shopping street or introducing new characters into a virtual room during a role-play scenario. The setup of the VR technology is illustrated in Fig 3. This technology enables the development of customized VR scenarios tailored to diverse client needs and treatment goals. Screenshots of various virtual environments of the VR application are displayed in Fig 4.

**Fig 3.**
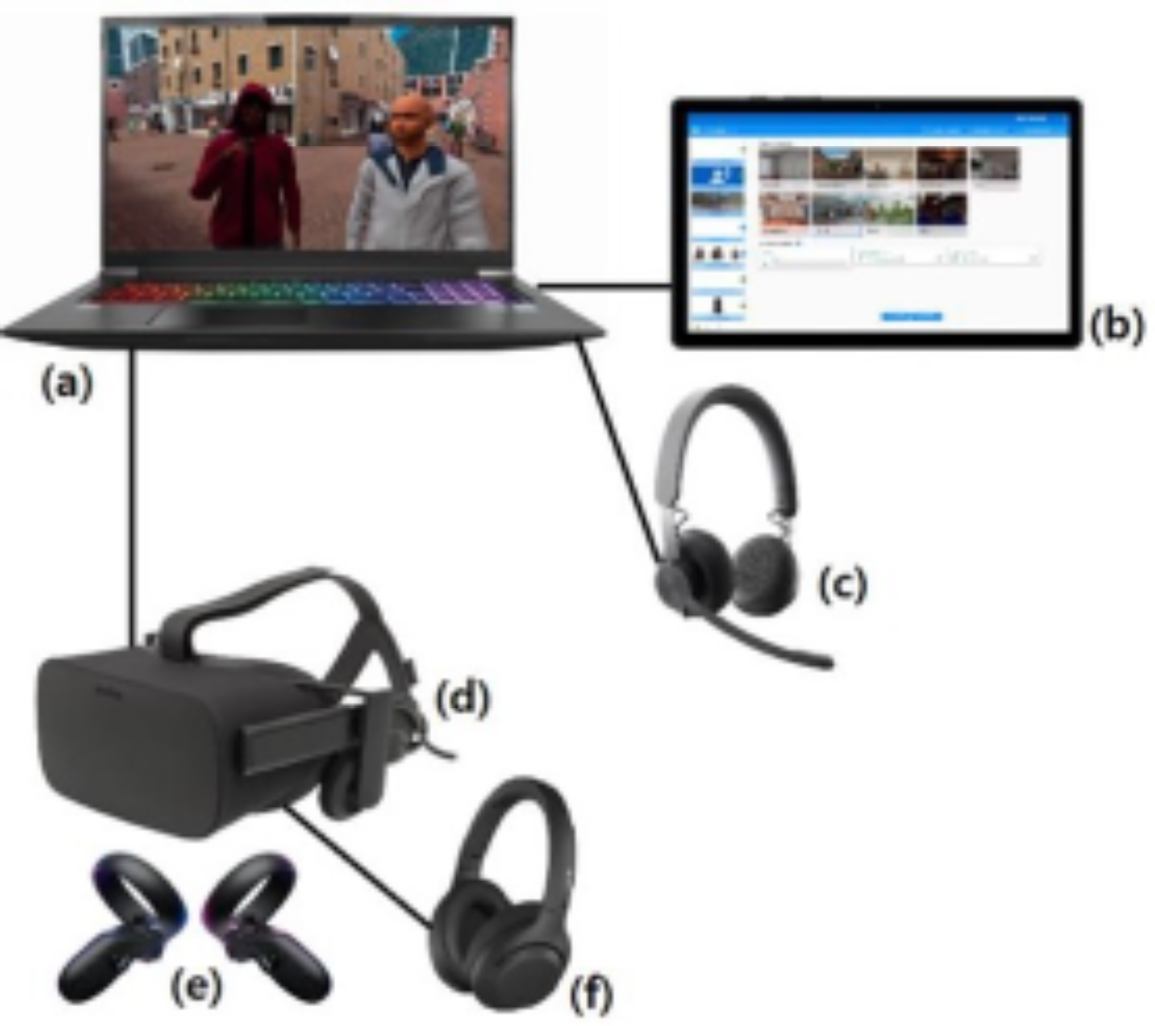
Setup of VR technology consisting of a laptop (a), a tablet with the dashboard (b), a voice-morphing microphone (c), a VR head-mounted display (d), VR controllers (e), and headphones (f)

**Fig 4.**
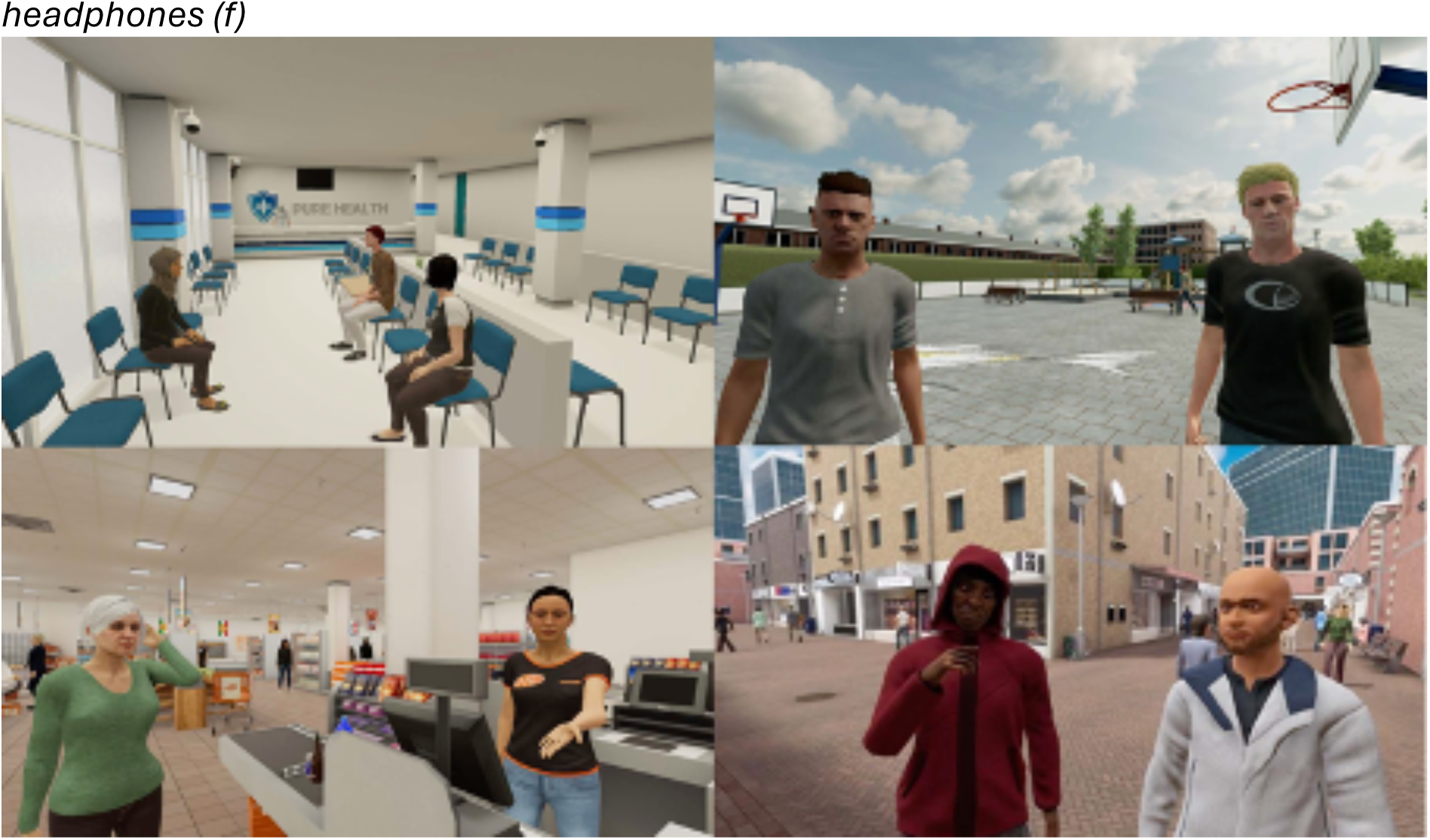
Screenshots of a selection of virtual environments in which the patient can walk around or perform a role-play (©CleVR)

### Implementation strategy: VR Training Program

Initially, the implementation process of the VR technology described in this paper started in 2019, with the decision of management to buy two VR sets of CleVR. This initial implementation pilot is described in detail in the case study of Kip and colleagues [19]. The VR training program that is described in the current study is a new implementation strategy that was put into practice based on the lessons learned from that previous pilot [19]. To develop a training that has a solid foundation to help integrate VR in practice, a systematic approach was used to address hindering and facilitating aspects related to the implementation of VR found in previous studies (Kip et al., 2023; Kouijzer et al., 2023). These implementation factors identified in previous studies were categorized according to the domains of the Consolidated Framework for Implementation Research [36], which provides a broad perspective on implementation at multiple levels (e.g. inner setting, individuals, and intervention characteristics). In contrast, the current study applies the TDF and COM-B model to specifically examine how individual therapists’ behaviors evolve during VR implementation. This distinction allows us to build on prior findings while focusing on the mechanisms underlying therapist behavior change.

In developing the training, the previously identified implementation factors are linked to specific training activities and sessions in which these activities take place (Table 1). The training program was developed in a co-creation process with a multidisciplinary team consisting of two researchers, two policy officers, and one therapist. The translation of implementation factors into concrete training activities was done through an iterative process, in which the team ensured that each barrier was addressed through targeted exercises, discussions, or hands-on practice. Additionally, the training design was informed by existing structured training programs for other treatments, such as cognitive behavioral therapy and EMDR, which emphasize the importance of thorough education, supervision, and skill development before clinical application [37, 38].

**Table 1.** The set-up of the VR training program based on identified implementation factors by [1S].

| **Implementation factors related to the training [19]** | **Implementation objectives targeted in the training [19]** | **Training activity** | **Training session** |
| --- | --- | --- | --- |
| **CFIR domain: Characteristics of individuals - therapists** |  |  |  |
| Investing time and effort | Therapists have sufficient intrinsic motivation to invest time and effort to work with VR. | Discussion on VR's added value during group discussions. | Session 1 |
| Integration of VR in routines | Therapists are able to smoothly integrate VR in their treatment sessions. | Practice setting up VR equipment and initiating sessions.<br>Discussing the integration of VR in treatment workflow. | Sessions 1-5 |
| Knowledge and skills | Therapists experience that they have sufficient technical and therapeutic knowledge and skills to use VR with their patients. | Hands-on practice operating VR hardware, navigating modules, case-based exercises. | Sessions 1-5 |
| Attitude towards technology | Therapists have sufficient knowledge about the current state of affairs regarding scientific research on VR. | Overview of VR studies and case examples in a presentation by a researcher. | Session 1 |
| Topic of conversation among colleagues | Therapists regularly discuss VR with trained and untrained colleagues to share experiences and/or information. | Reflection and exchange of experiences in peer groups. | Throughout and after the training, in addition to sessions. |
| Experienced benefits/added value | Therapists are aware of the place of VR in the mission and vision of the organization. | Discussion of organizational goals and VR alignment. | Session 1 and 6 |
| **CFIR domain: Inner setting** |  |  |  |
| Introduction of VR to therapists | The organization structurally organizes a broad range of activities to keep trained and untrained therapists informed about VR in general. | Presentation and demonstration of VR and their potential uses within therapy. | Session 1 and 6 |
| Content-related support for therapists | There is a clear overview of available resources and support systems set up by the organization. | Dissemination of learning materials such as instructional guides, tips and tricks for VR usage and a Teams environment. | Session 1 and homework assignments. |
| Necessary preconditions for usage | The organization ensures that the practical requirements of therapists are met to ensure that all practical barriers to use VR are removed. | Sufficient availability of VR sets and treatment rooms. | Before the training. |

The main goal of the VR training program was to equip participating therapists with the knowledge and skills necessary to proficiently use VR in therapeutic practice. The training focused on providing participants with a comprehensive understanding of handling VR equipment, navigating VR environments, recognizing therapeutic situations where VR can enhance treatment outcomes, and integrating VR into daily workflow. This included selecting appropriate VR modules based on treatment goals, preparing and setting up the VR system within a session, guiding patients through VR exercises, addressing potential technical issues, and debriefing patients afterward to integrate VR experiences into therapeutic sessions. The training program consisted of six sessions, starting with an introduction to VR technology and its therapeutic potential. In the following sessions, therapists practiced navigating VR environments in small groups, taking on the roles of practitioner, patient, and observer. Later sessions focused on role-playing interactions with VR characters to simulate real-world therapeutic scenarios.

The final session emphasized reflective learning through a video assignment, where participants recorded a VR session with a patient and provided peer feedback to each other’s recordings. Throughout the program, therapists completed homework assignments and practiced VR usage with patients in their treatment sessions. The training was delivered by two trainers: a policy officer, who ensured the right organizational conditions and provided technical support, and a therapist, who guided participants in the clinical application of VR.

### Participants

Interviews were conducted with eleven therapists who all participated in the VR training. Inclusion criteria for therapists were that they 1) currently worked for the mental healthcare organization, 2) were involved in any type of mental healthcare treatment with in- or outpatients, and 3) they participated in the VR training program.

The recruitment of participants was carried out by the coordinator and trainer of the VR training program, an innovation policy officer. All employees of the mental healthcare organization who matched the first two inclusion criteria received a message via the employee portal and/or were informed by their manager about the possibility of signing up for the VR training program. The participants had to register themselves via email to the coordinating trainer of the program. The participants of the training were approached by the researcher (MK) via email with the request to participate in the study related to the VR training. In total, eleven out of the twelve participants of the training agreed to participate in the study (Table 2). One participant mentioned not being interested in participation. In the first round of interviews, all eleven participants were interviewed. After the first training session, one participant could not continue with the training due to scheduling issues, another participant could not continue due to illness, and the last participant had to stop due to changing organizations. These participants did not complete the second and third interview rounds. After two training sessions, another participant had to stop participating due to illness. In total, seven participants completed the VR training program and participated in all interviews.

**Table 2.** Characteristics of participants.

| Participant # | Function | Work experience<br>mental healthcare<br>in years | Patient group | Participated<br>in interview # |
| --- | --- | --- | --- | --- |
| 1 | Psychologist in training to become behavioral therapist | 6 | Complex trauma | 1, 2, 3 |
| 2 | Cognitive behavioral therapist & System therapist | 19 | Forensic | 1, 2, 3 |
| 3 | Psychologist | 4 | Anxiety, mood, and personality disorders | 1, 2, 3 |
| 4 | Social psychiatric nurse | 24 | Bipolar and psychotic disorders | 1, 2, 3 |
| 5 | Social pedagogical counselor | 6 | Anxiety, mood, and personality disorders | 1, 2, 3 |
| 6 | Psychomotor therapist | 7 | Complex trauma & Anxiety and mood disorders | 1, 2, 3 |
| 7 | Social worker & Social psychiatric nurse | 1 | Youth | 1 |
| 8 | Psychologist | 2 | General | 1, 2 |
| 9 | Clinical psychologist & Cognitive behavioral therapist | 20 | Youth | 1 |
| 10 | Nurse practitioner & Lead practitioner | 33 | Addiction | 1 |
| 11 | Social worker | 23 | Youth | 1, 2, 3 |

### Materials and Procedure

Data were collected by one researcher (MK) through in-depth, semi-structured interviews. Three interview schemes were developed to gain in-depth insights into the expectations and experiences of the therapists with the VR training program. The first draft of the interview protocols was improved based on a pilot test with a therapist - who did not take part in the VR training - and in consultation with another researcher (HK). After the participants were informed about the study, they had the opportunity to ask questions. They signed the informed consent form before the first interview took place.

The first round of interviews took place in October 2023. The participants were asked several introductory questions about their current function, years of work experience in mental healthcare, and experience with VR. Second, their expectations regarding the VR training program were explored, focusing on activities, possible barriers, and advantages of the program. Lastly, they were asked to formulate concrete personal goals regarding their VR usage over the next 6 months.

The second round of interviews took place in January 2024, directly after the training was concluded. The participants were asked about their experiences with the VR training program, experienced positive aspects, and points of improvement. Furthermore, questions were asked about the match between their expectations before the training and their experiences after. Finally, it was discussed to what extent participants achieved their personal goals for VR use.

The third round of follow-up interviews with therapists took place in April 2024, three months after the VR training was concluded. First, the participants were asked about their experience in the past three months of working with VR in practice. Second, their personal goals were evaluated again. Third, the added value of VR and additional points of improvement in the implementation process of VR technology were discussed. Finally, they were asked about expectations for future VR use in practice.

All interviews took place at the location of preference of the participants. In all cases, participants preferred to conduct the interviews online, via Microsoft Teams, a videoconferencing program. All interviews were conducted by one researcher (MK) and were audio recorded. On average, the interviews took 27 minutes, ranging from 15 to 37 minutes. The interview protocols are presented in Appendix A.

### Data Analysis

The audio recordings of the interviews were transcribed verbatim by one researcher (MK). First, all transcripts were read to familiarize with the data and identify meaningful fragments based on the objectives of the study. A deductive coding scheme was developed by one researcher (MK), while another researcher (HK) remained consistently engaged in the process, providing continuous feedback and oversight. First, the fragments were categorized deductively based on the categories of the COM-B model and the domains of the TDF. This approach enabled us to systematically map the data onto established determinants of implementation behavior. Second, we categorized experiences per domain as positive or negative to identify key strengths and areas for improvement. Negative evaluations were analyzed alongside suggested improvements, providing actionable insights for effective implementation. This structured, top-down analysis ensured a comprehensive evaluation of the VR training program.

In the analysis, we also paid attention to how these determinants evolved across the three time points, allowing us to observe shifts in capability, motivation, and opportunity over time as therapists gained more experience with VR. These changes were visualized using bar charts, which depict the number of both positive and negative evaluations per determinant and across each interview round, providing a clear overview of the trends and shifts in therapist perspectives over time. The coding process was first discussed by two researchers, after which one researcher (MK) coded all fragments. Next, the second researcher (LK) checked 10% of the coded fragments. The researchers’ opinions differed on only 16% of the codes, relating to 84% consensus. Based on the differences, minor adjustments were made to the allocation of codes to ensure consistency and accuracy.

## Results

In this section, the findings of the current study on determinants influencing the implementation of VR in mental healthcare are presented. The findings from each interview round are presented in Tables 3, 4 and 5, corresponding to the pre-training, post-training, and follow-up interviews, respectively. The determinants are derived from the combined COM-B model [39] and the Theoretical Domains Framework [40], as illustrated in Fig 1. The evolution of determinants over time is visualized in Figs 5, 6, and 7.

**Fig. 5.**
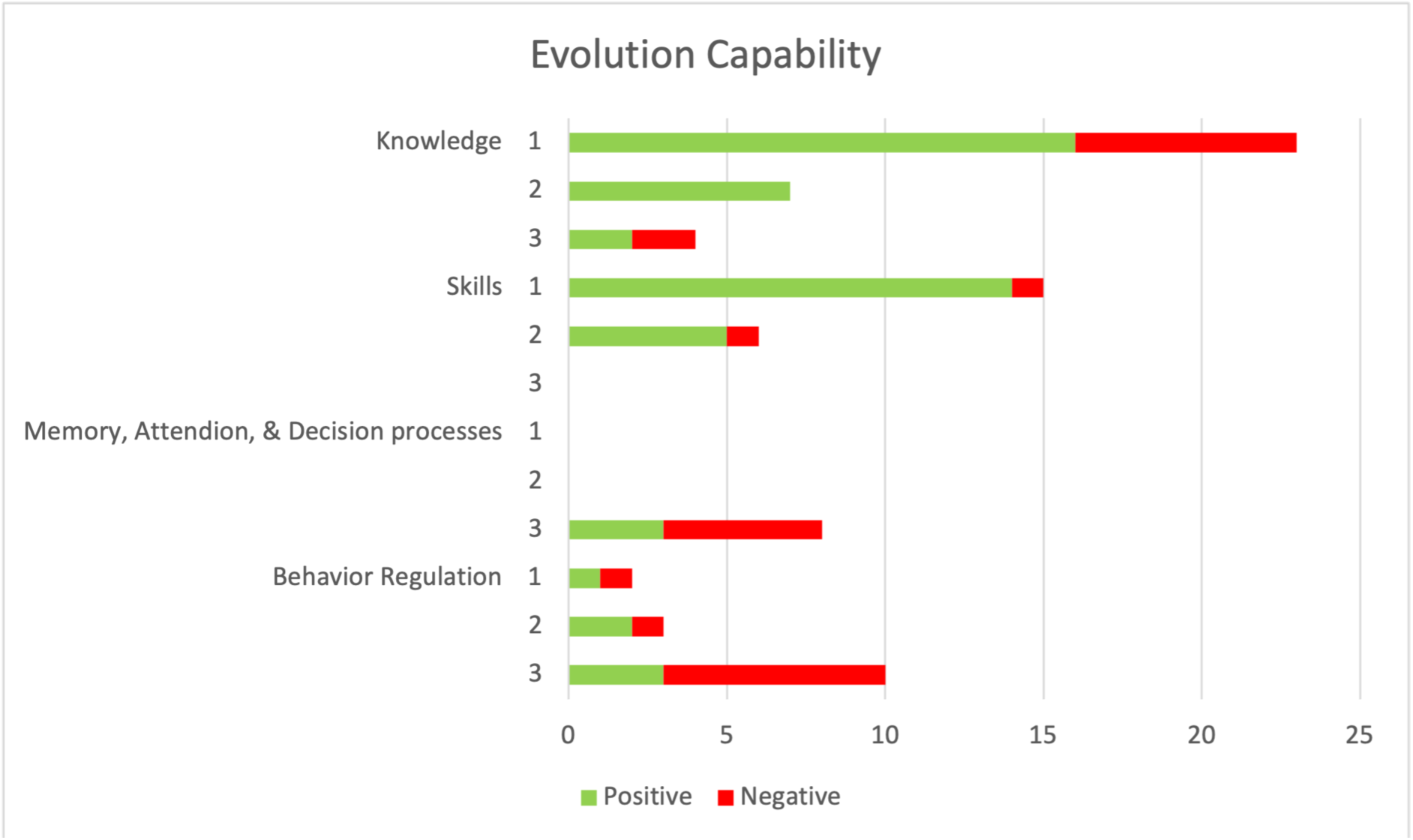
The Evolution of Determinants related to Capability that influenced VR Implementation over time.

**Table 3.** Determinants related to Capability that influence VR Implementation according to Therapists.

| Domain & Definition | Interview round | Ntot<sup>a</sup> | N+<sup>b</sup> | N-<sup>c</sup> | Quote + | Quote - |
| --- | --- | --- | --- | --- | --- | --- |
| **Knowledge**<br>*'Therapists' awareness and familiarity with using VR.'* | 1 | 23 | 16 | 7 | "I know there are colleagues who haven't had such extensive training. (...) I don't think that's okay because I believe you need to have sufficient knowledge before using it." (p2) | "Yes, of course, you need to know how it [VR] works, but right now I have no idea how to go about it." (p4) |
|  | 2 | 7 | 7 | - | "I appreciated that some background information was shared. I usually prefer practicing hands-on, but it's definitely good to know the theory behind it as well, and you can certainly find that in the training." (p6) | - |
|  | 3 | 4 | 2 | 2 | "I do have the knowledge to use it. If I were to start doing it now, it would come back to me right away." (p6) | "I notice that I still sometimes find it difficult to know when it's appropriate to use it or not. (...) But I think it's simply a matter of gaining experience." (p5) |
| **Skills**<br>*'Therapists' ability or proficiency in VR use acquired through practice.'* | 1 | 15 | 14 | 1 | "I expect to become skilled in operating the equipment, to learn about its possibilities and applications in treatment, and to gain insight into suitable case studies." (p11) | "At the moment, I don't feel well-prepared to use VR; I think I still need to get comfortable with the technical aspects." (p2) |
|  | 2 | 6 | 5 | 1 | "I do think I could use VR in practice now. I believe I can do it." (p3) | "I didn't learn how to apply VR in a group setting. That part remained really vague in the training. How do you actually do that in practice?" (p5) |
|  | 3 | - | - | - | - | - |
| **Memory, Attention, & Decision processes** | 1 | - | - | - | - | - |
| *'The extent to which using VR is part of therapists' regular practice.'* | 2 | - | - | - | - | - |
|  | 3 | 8 | 3 | 5 | "I do think about VR as a treatment option when I start working with someone. During the introduction, I consider whether it would be a good fit or not." (p4) | "It's still not an automatic part of my thinking when I assess someone. I'm just not focused on it at that moment. It's not part of my automatic response." (p3) |
| **Behavior Regulation** | 1 | 2 | 1 | 1 | "Yes, I would appreciate having peer supervision groups and time with colleagues. Also because I know that I wouldn't keep this in focus on my own. So I find that very beneficial." (p5) | "If I start using VR, it requires quite a bit of planning and organization." (p1) |
| *'Therapists' ability to self-monitor and action plan to use VR.'* | 2 | 3 | 2 | 1 | "My learning style is that I like to have everything clear in advance. (...) Some people are more like, 'I'll just do it and see what happens.' I'm definitely more on the other side." (p8) | "Often, you plan in this work so shortly in advance, and then you don't have much time beforehand to consider using VR." (p2) |
|  | 3 | 10 | 3 | 7 | "I do try to practice as much as I can on my own. I regularly make an effort to sit down with it so I can keep my knowledge up to date on how to use it. Otherwise, it will be even harder when I need to use it with patients." (p5) | "The reason I use VR less now is that you really need to have a clear plan in advance and reserve the equipment. So all those preparations don't really align with my current flexible working style." (p6) |

<sup>a</sup> The number of times the determinant is mentioned in that specific interview round (Ntot)
<sup>b</sup> The number of times the determinant is mentioned as positive point in that interview round (N+)
<sup>c</sup> The number of times the determinant is mentioned as point of improvement in that interview round (N-)

**Table 4.**
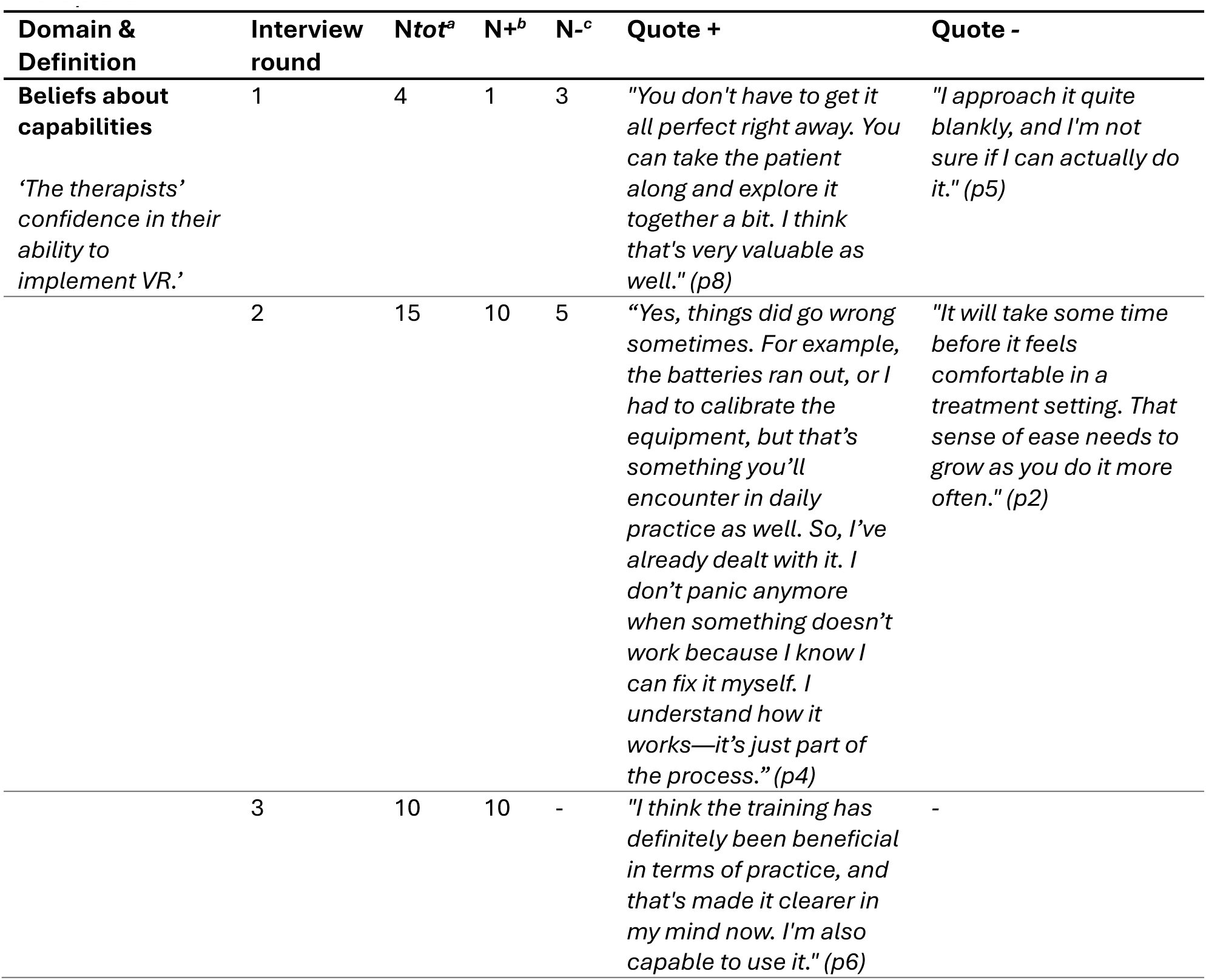
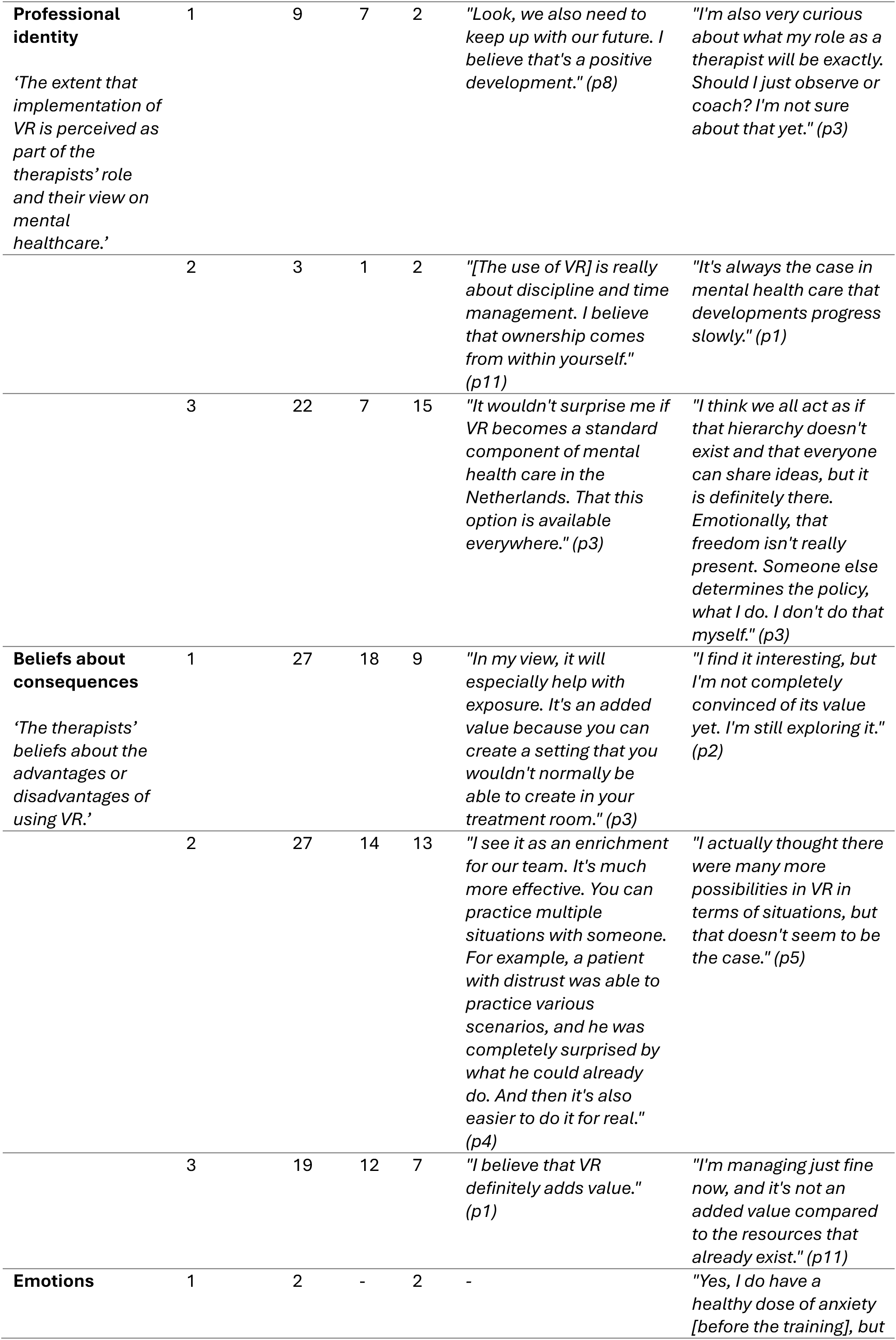
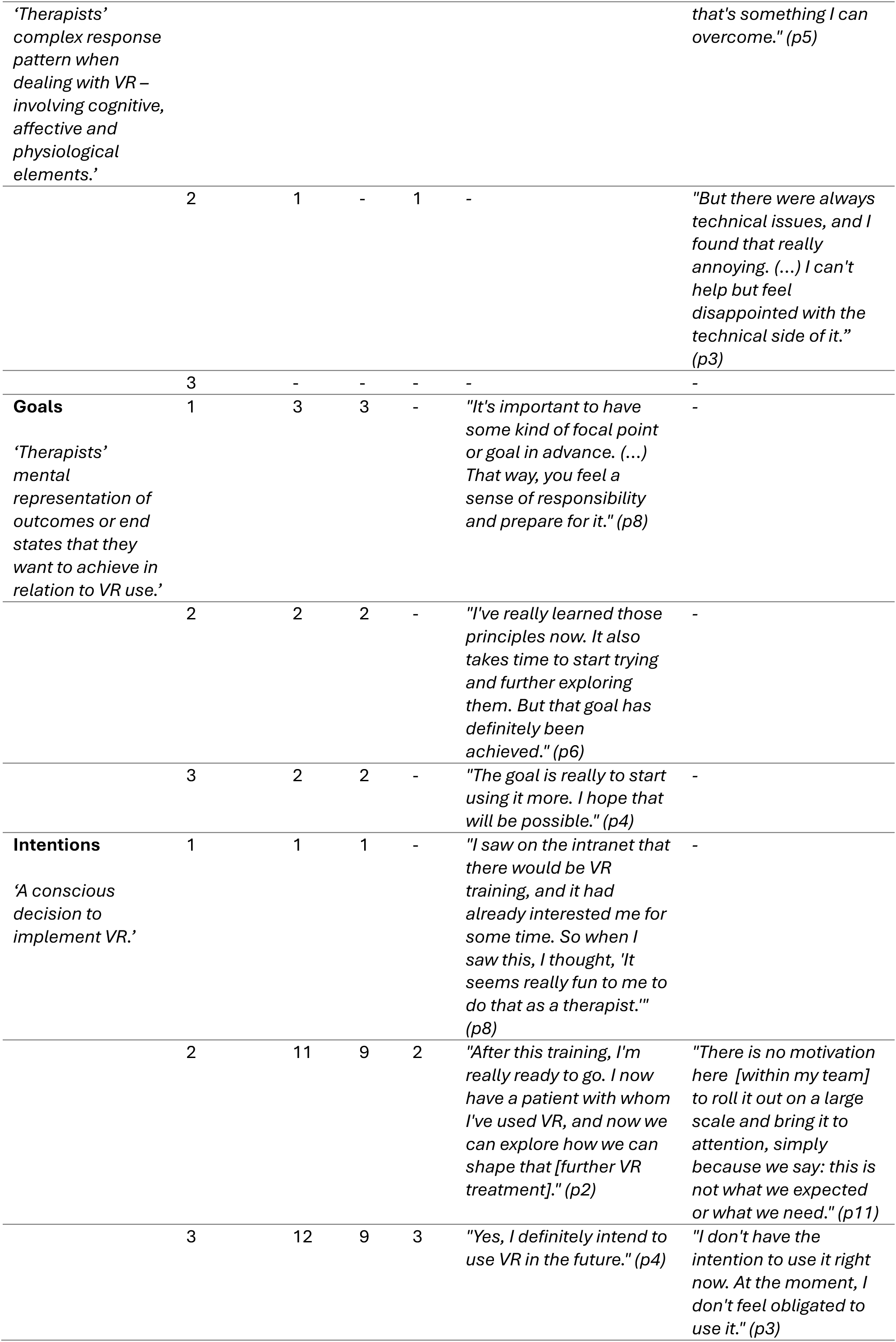
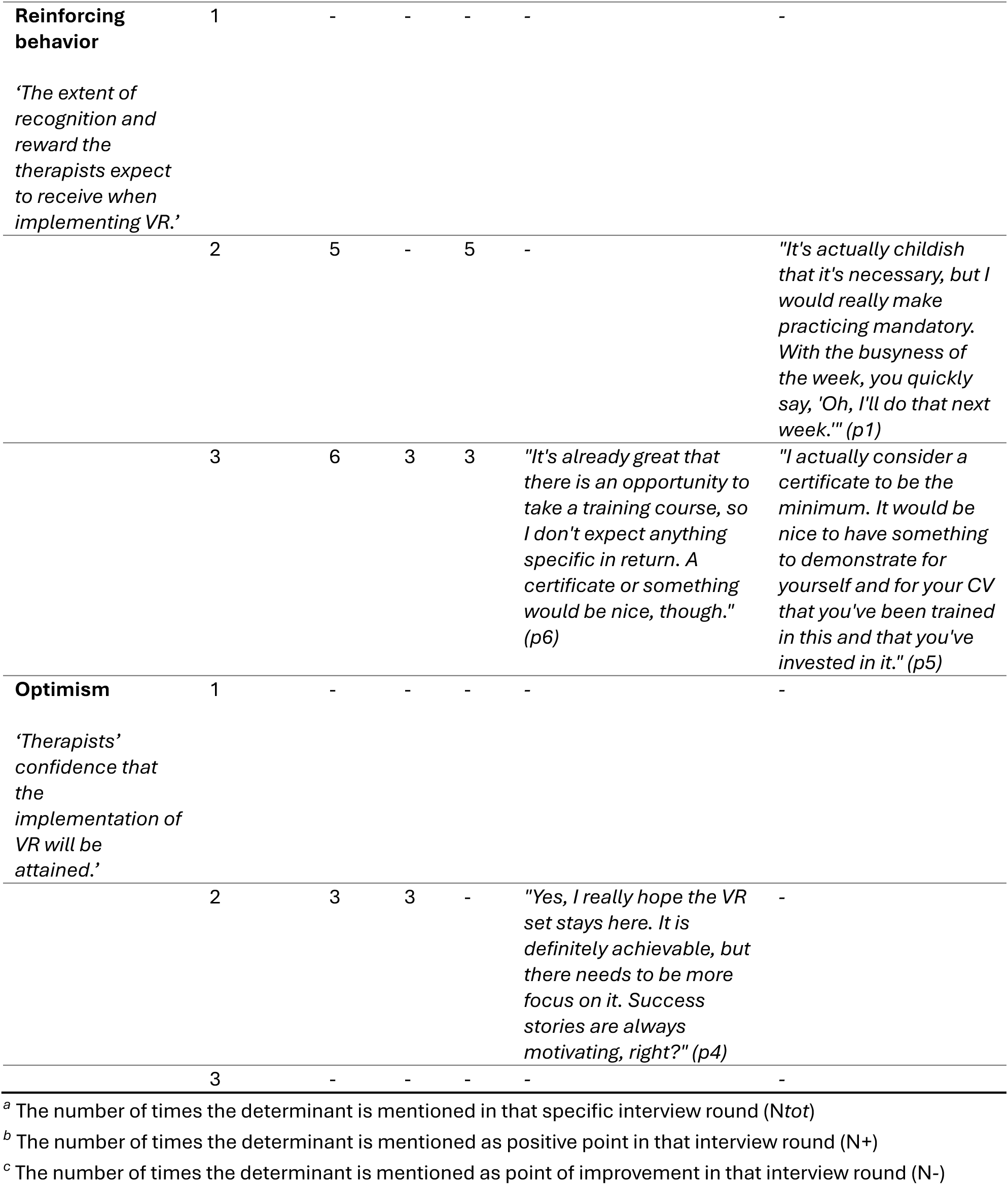
Determinants related to Motivation that influence VR Implementation according to Therapists.

### Capability

The ***capability*** category explores determinants that relate to the therapists’ psychological and physical capacity to implement VR in their treatment practice [39]. In Table 3, all determinants within the capability category that influence the implementation of VR technology in mental healthcare, according to therapists, are described. Fig 5 illustrates how these determinants evolved over time.

In total, ***knowledge*** was the most frequently mentioned determinant across all interview rounds, with positive remarks about the extensive training program increasing their knowledge, but also negative remarks about the overall difficulty of applying VR in treatment. A noteworthy shift was observed in therapists’ perceptions regarding their knowledge related to VR use. Initially, therapists highlighted their lack of knowledge of VR, particularly in its application, indications, contraindications, and integration into treatment. During the training, therapists gained knowledge, and three months after the training concluded, all therapists mentioned retained knowledge of VR usage in treatment. Some therapists mentioned needing more practice to apply the knowledge in practice. All therapists indicated that the training had addressed initial gaps, fostering confidence in their ability to use VR in clinical practice.

Therapists mentioned that their ***skills*** related to VR use also improved. Therapists expressed a high level of proficiency in using VR and integrating it into their treatment after the training, with few negative comments on their competence. This positive shift suggests that the therapists felt they had acquired the necessary skills to incorporate VR into their practice. Therapists mentioned that they retained their VR skills over time.

Interestingly, in later interviews, therapists hardly mentioned skills as a topic for discussion. While initially perceived as a key learning objective, the focus on skill development diminished after the training. At the same time, other factors received more attention as the interviews progressed, indicating that other determinants became more pressing.

Regarding ***memory, attention, and decision processes*** and ***behavioral regulation***, therapists acknowledged three months after the training that VR had not yet become a standard treatment option. Despite their increased confidence in using VR, they did not automatically think of it as a standard option when creating their treatment plans. Many therapists still reverted to more familiar treatment methods, highlighting the lack of automatic integration of VR into their decision-making processes. The structured planning and coordination for VR use was mentioned as a challenge, as therapists’ treatment routines typically involved spontaneous adaptations based on patient needs, rather than the scheduling that VR requires in practice. Interestingly, this topic had not been discussed in the first two interviews, possibly indicating that it had not been a priority in their initial thinking.

Overall, therapists felt confident in their capacity to use and integrate VR into their clinical practice, indicating the retention of knowledge and skills over time. Despite this capability, therapists mentioned that they did not use VR as often as they would have thought when they started the training.

### Motivation

The ***motivation*** category explores determinants that relate to therapists’ internal process that initiates, directs, and sustains goal-oriented behavior. It includes not just goals and intentions, but also habitual processes and emotional responses [39]. In Table 4, all determinants are presented that played a role in shaping therapists’ motivation to implement VR into mental healthcare. Fig 6 illustrates how these determinants evolved over time.

**Fig 6.**
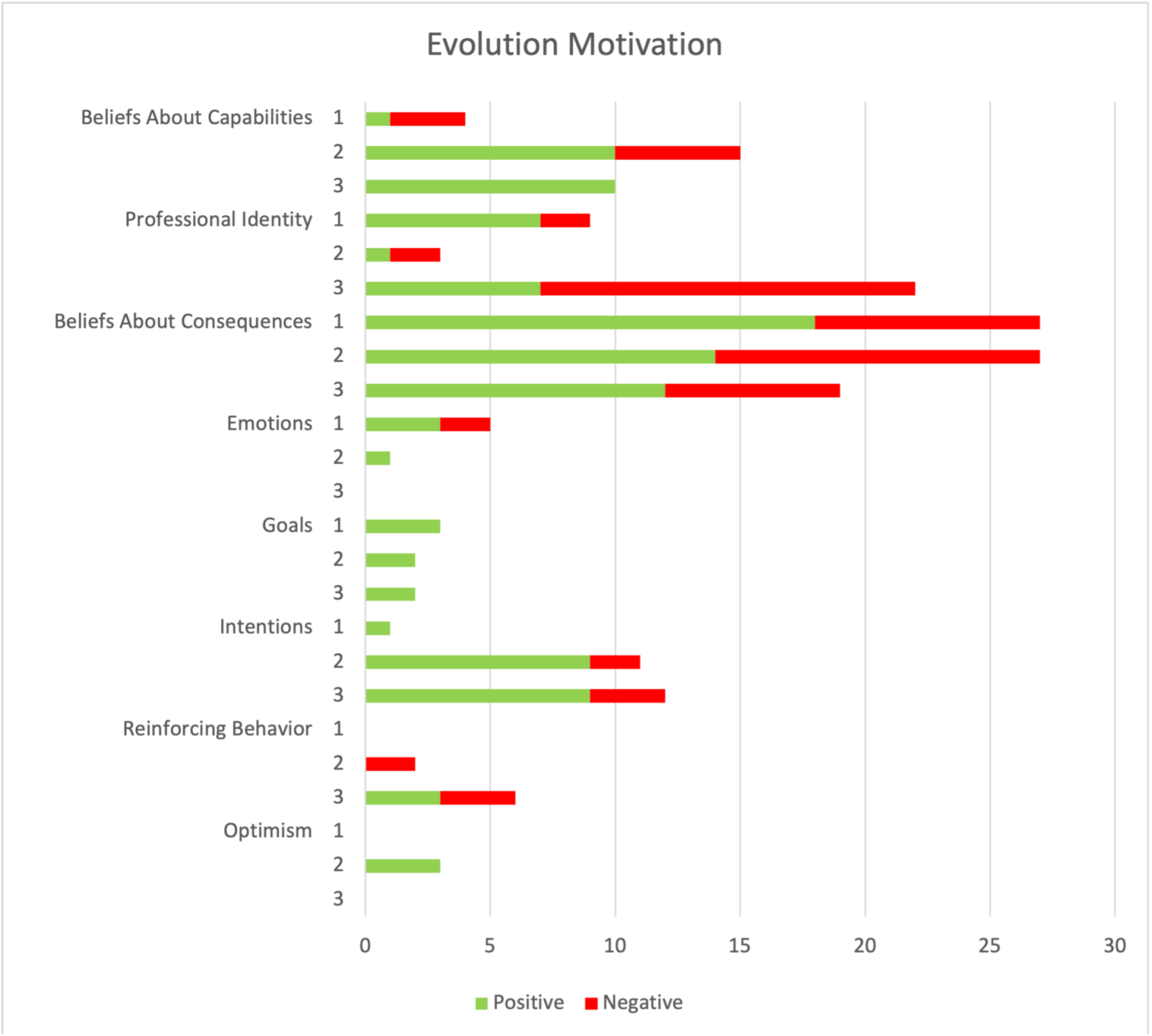
The Evolution of Determinants related to Motivation that influenced VR Implementation over time.

At the outset, many therapists expressed reservations in their ***beliefs about capability*** to integrate VR into their practice. However, throughout the training, confidence grew, and all therapists reported a belief in their capability to use VR effectively. The determinant ***beliefs about consequences*** of VR use emerged as a key factor, with therapists recognizing the potential of VR for exposure therapy and appreciating its benefits, such as providing a controllable and safe environment for various scenarios. A therapist even suggested that VR could be more effective than traditional treatments, especially in helping patients transfer coping strategies to real-life situations. Despite these positive beliefs, concerns remained about the lack of realism in some virtual scenarios and the challenges in finding suitable patients for VR treatment. Most of their patients were able to follow exposure therapy outside the clinic, making the use of VR unnecessary. Other patients required medication for more pressing symptoms before engaging in VR treatment.

The ***professional identity*** of therapists became more prominent as the training progressed. Therapists increasingly saw the potential for VR to become a standard treatment option in mental healthcare. However, some reported feeling constrained by hierarchical team dynamics that hindered their willingness to stimulate VR use after the training concluded. The perception of limited autonomy within their teams restricted their motivation to advocate for the continued use of VR.

Furthermore, the therapists’ attention to ***reinforcing behavior*** regarding the use of VR increased over time. Most therapists advocated for stricter requirements for participating in VR training and receiving certification to encourage responsible VR use. They believed that setting higher entry requirements would enhance motivation, accountability, and discipline among those wishing to integrate it into their therapeutic practice.

In the interviews, emotions, goals, intentions, and optimism were mentioned less frequently. Relating to ***emotions***, therapists mentioned some anxiety about using VR, particularly about technical issues that may occur, but these emotions decreased over time. Regarding ***goals***, therapists highlighted the importance of having a clear objective when using VR, with some aiming to integrate it more into their practice. ***Intentions*** were expressed mostly positively, especially after training, with therapists feeling ready to implement VR, though some struggled with team-wide motivation. Lastly, ***optimism*** for VR’s future in treatment was expressed, but this was less frequently discussed.

### Opportunity

The ***opportunity*** category explores determinants that relate to all the factors that lie outside the therapist that make the implementation of VR possible or hinder it [39]. In Table 5 both determinants within the opportunity category are presented. Fig 7 illustrates how these determinants evolve over time.

**Table 5.**
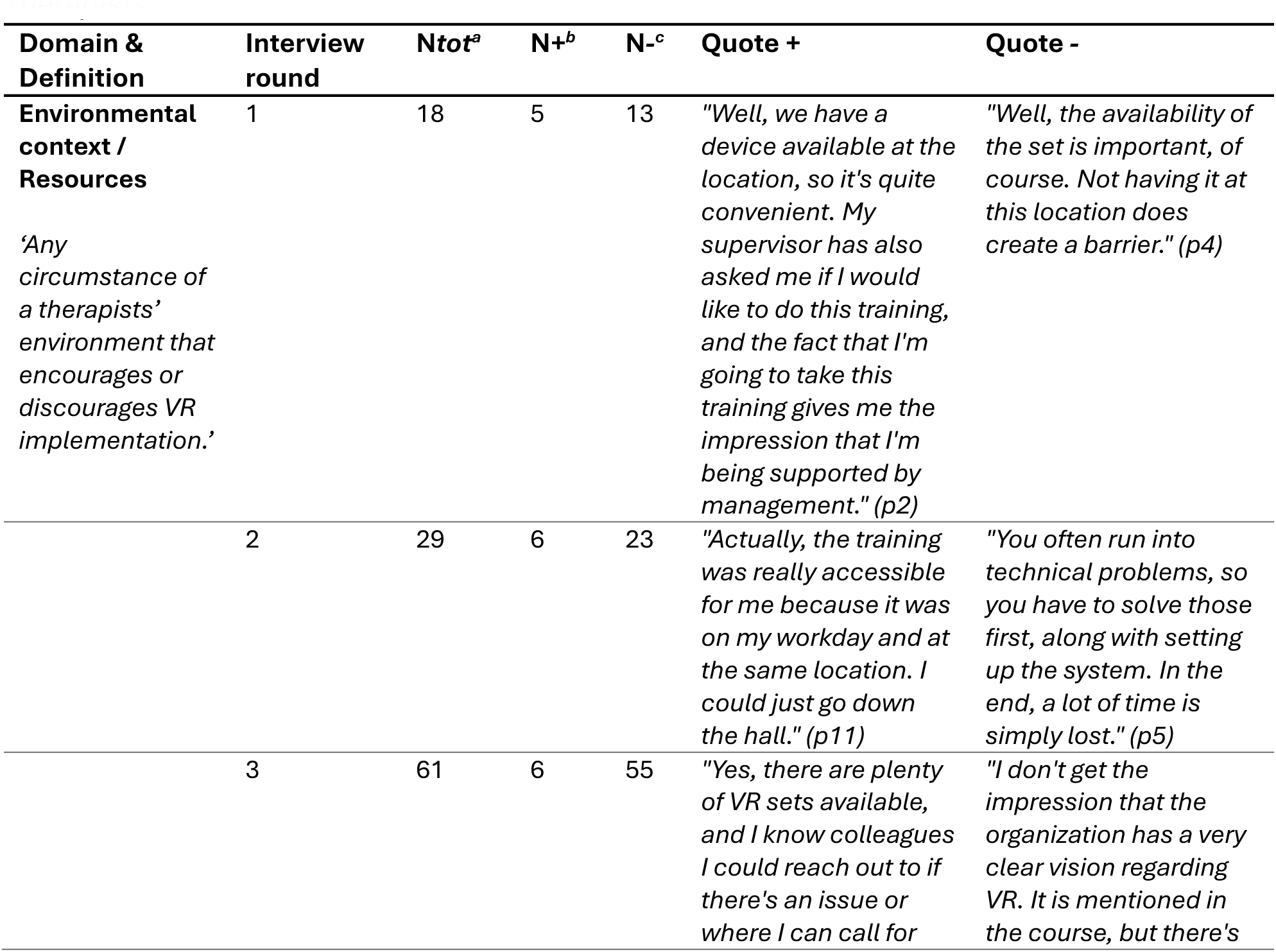
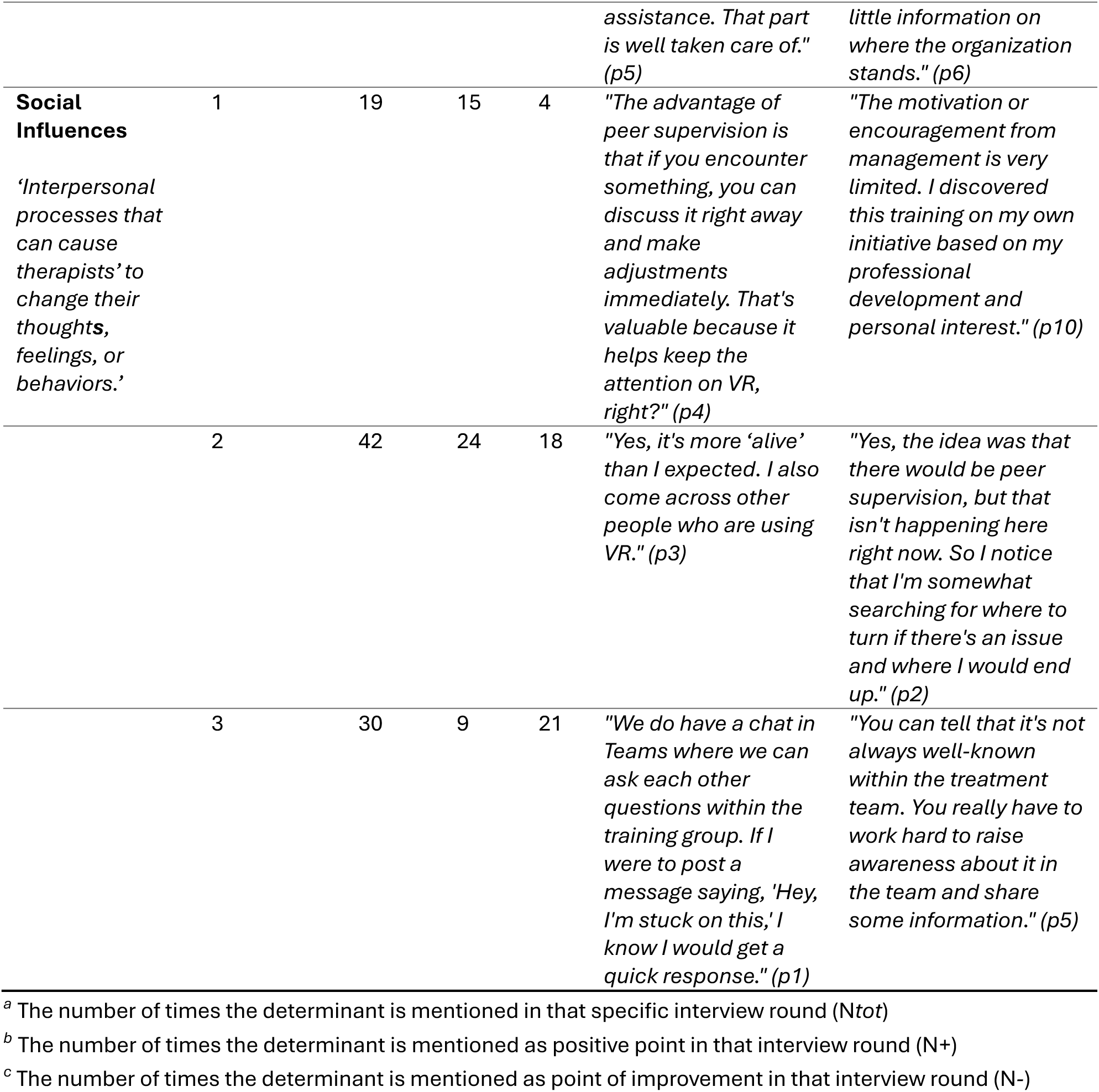
Determinants related to Opportunity that influence VR Implementation according to Therapists.

**Fig 7.**
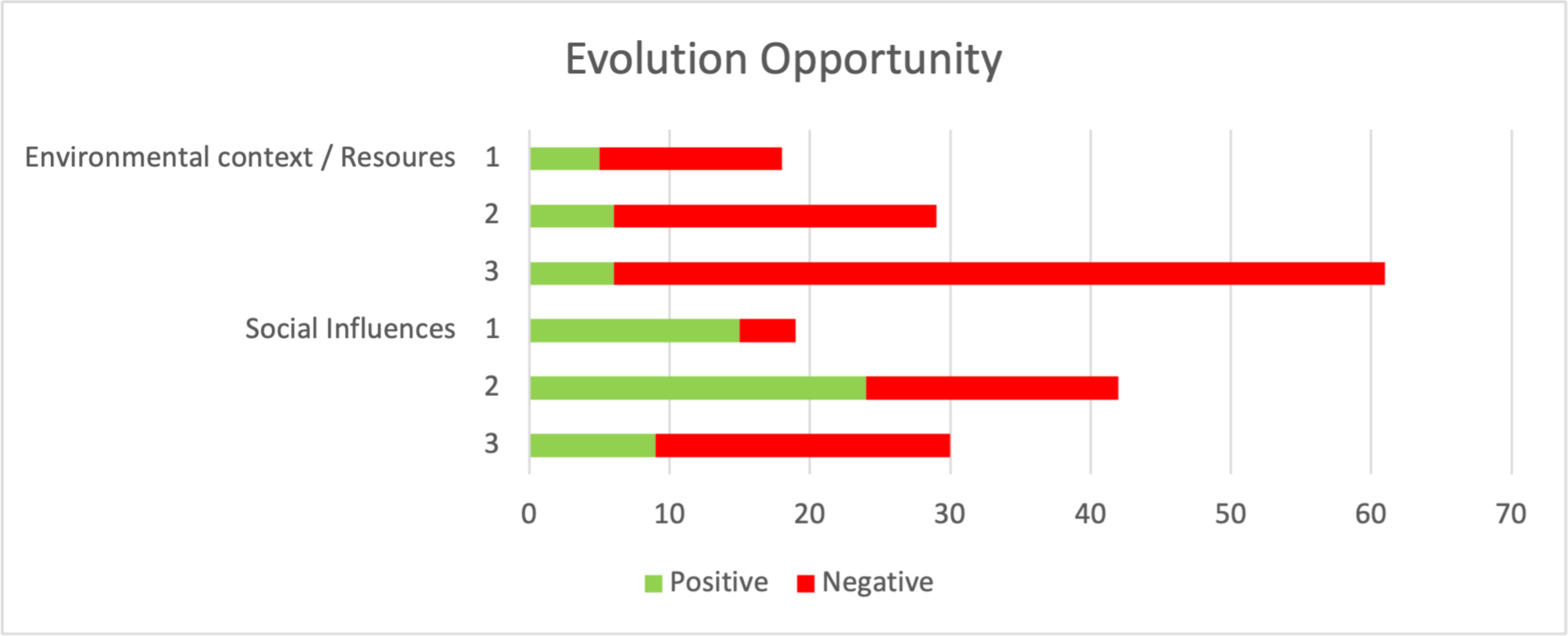
The Evolution of Determinants related to Opportunity that influenced VR Implementation over time.

Both determinants, ***environmental context/resources*** and ***social influences,*** emerged as critical in understanding therapists’ experiences and attitudes toward the implementation of VR. Initially, therapists identified the availability of VR sets and training as facilitating factors. However, as the study progressed, these were overshadowed by a growing sense of frustration due to an increasing number of challenges, such as technical difficulties with the VR equipment and issues with reserving the necessary resources. Therapists reported that these complications, combined with overwhelming workloads, led to an increasing sense of dissatisfaction and left little room to explore and integrate VR into their treatment practices.

A significant lack of organizational vision and sustained support after the training regarding using VR in treatment was also noted. Therapists felt that management did not prioritize VR and provided limited support or communication after the training ended.

This clear lack of direction from the organization created an environment where therapists felt largely unsupported in their attempts to integrate VR into their treatments. By the final round of interviews, many therapists voiced the need for sustained organizational support and a robust vision for integrating VR and innovations in general into mental healthcare.

Regarding ***social influences***, therapists initially experienced a sense of enthusiasm and community among colleagues, but this supportive atmosphere diminished over time. Therapists reported feelings of isolation and disappointment stemming from a lack of ongoing encouragement from colleagues, team leaders, and management. Many expressed that the absence of engagement and motivation from leadership further exacerbated these feelings, decreasing their drive to use VR. Three months after the training concluded, therapists indicated a need for a more active role from management and for increased awareness and advocacy within their teams to foster a supportive environment.

Overall, these two determinants were more frequently discussed than other determinants in the TDF framework. Issues ranged from broader organizational challenges to specific team dynamics and individual colleague interactions.

## Discussion

### Principal findings

This longitudinal evaluation explored how a specific implementation strategy – a VR training program – impacts therapists’ VR use and implementation behavior. The evaluated behavior determinants are derived from the TDF and COM-B model [39]. The key findings are related to the capability, motivation, and opportunity of therapists to implement VR. Regarding capability, therapists improved and retained their self-reported knowledge and skills in using VR during and after the training. Despite these improvements and the positive intention to use VR, it was not structurally integrated into treatment once the training had been concluded. This can largely be attributed to determinants related to opportunity that became apparent over time, such as a lack of organizational vision and sustained support from management, as well as the technical complexity of VR technology. Notably, these barriers only became apparent after therapists had gained sufficient knowledge and skills to consider long-term implementation, suggesting that capability development is a prerequisite for identifying and addressing broader implementation challenges. This highlights not only the importance of a longitudinal evaluation but also the need for a phased implementation approach, where different implementation strategies are deployed at different stages. While the capability and motivation to use VR improved, organizational and technical barriers emerged later in the process, emphasizing that implementation requires ongoing, adaptive strategies rather than one-time intervention.

### Comparisons to prior work & Implications for practice

The VR training program effectively supported knowledge acquisition and positively impacted various other determinants, including the development of skills necessary for VR use in practice. Importantly, this extended beyond technical proficiency – therapists also reported feeling capable of integrating VR into their treatment and applying therapeutic skills during VR sessions with patients. These findings align with research emphasizing that successful implementation of new technologies requires more than just technical training; professional development should also focus on broader topics such as privacy, ethical and legal considerations, clinical competencies, and administrative tasks [41]. However, despite these self-reported improvements in knowledge and skills, VR was not consistently integrated into daily treatment practice after the training concluded. Therapists tended to revert to familiar treatment methods rather than incorporate VR as a standard part of their treatment. This is often seen in VR implementation, as therapists tend to feel more comfortable and confident in the effectiveness of their familiar methods and these methods are perceived as more straightforward and reliable [42–44]. Some other key barriers to implementation were the high perceived workload and time constraints, which made it difficult for therapists to reserve and set up VR equipment in time for treatment. This highlights the importance of considering context-related obstacles in implementation efforts. This is also emphasized in a study on the implementation of e-mental health applications in Belgian psychiatric hospitals, which stated that high workload and lack of time often hinder the adoption of new technologies and innovations in mental healthcare [45].

Regarding motivation to use VR, therapists generally viewed VR as a valuable tool, particularly for interventions such as exposure therapy, where it could lower the threshold for real-life exposure. This positive attitude towards the technology reflects growing recognition of VR’s potential to enhance treatment. However, despite this enthusiasm, concerns about the perceived consequences of using VR in treatment, such as the lack of realism of VR scenarios and their suitability for patients, persisted even after the training. For instance, some therapists mentioned that patients found the VR environments too artificial, making it harder to immerse themselves in the experience. Others expressed concerns that the available scenarios may not adequately reflect the diverse needs and backgrounds of their patients, limiting their applicability. These concerns are in line with previous research showing that the alignment between technology and patients’ needs is a key factor in the acceptance and sustained use of VR in clinical practice [46]. When technology is tailored to specific patient groups, it is more likely to be perceived as useful, feasible, and engaging, ultimately enhancing treatment experiences and health outcomes [47, 48]. Although these concerns persisted, they did not necessarily prevent therapists from using VR. Some therapists noted that as they gained more experience with VR, their initial doubts about realism and suitability became less relevant, particularly when they observed positive patient outcomes. This suggests that while these concerns are valid, their true impact on clinical practice can only be determined through extended use over time. Indicating that implementation and evaluation go hand-in-hand and reinforcing the notion that implementation is an ongoing process, where some barriers can only be addressed once initial adoption has taken place [49, 50].

While the training improved therapists’ capability and generally increased motivation to use VR, several opportunity-related challenges hindered the sustained implementation of VR. This aligns with previous studies indicating that organizational factors are underemphasized in implementation efforts, often interrelated with technical and social dimensions, making them complex to address comprehensively [51]. One possible explanation is that many implementation strategies are designed in advance, making it difficult to fully anticipate challenges that only become apparent in later stages of the implementation process. Certain organizational barriers may not initially seem critical but can emerge as important obstacles once therapists attempt to integrate VR into routine practice. This highlights the dynamic nature of implementation and the need for adaptive strategies that evolve based on real-world experiences [50].

In addition, existing organizational theories often fall short of explaining the complexities of healthcare implementation [52]. As seen in this study, the combined TDF and COM-B model primarily focused on individual factors influencing behavior change, as reflected in the extensive list of determinants within the capability and motivation categories. However, the opportunity category is limited to only two determinants, which may be too minimal to provide a comprehensive and balanced understanding of how the organizational factors influenced the implementation process. Future research and implementation practice could integrate additional frameworks, such as the Consolidated Framework for Implementation Research (CFIR) [36] or the Non-adoption, Abandonment, and challenges to Scale-Up, Spread, and Sustainability framework (NASSS) [53]. These frameworks offer a more detailed examination of contextual determinants of implementation.

From this longitudinal evaluation, several implications for practice emerged. First, findings showed that enthusiasm for VR declined over time due to technical difficulties, limited availability of VR sets, high workload, and time constraints. Therapists frequently encountered software errors, found the setup process of the VR set complex, and often required technical support. These findings underscore the importance of ensuring that new technologies are not only functional but also seamlessly integrated into treatment without disturbing established workflows [54]. Despite the extensive training, these issues persisted, indicating that additional layers of support are essential for successful implementation. While training is important for building competence, it is not sufficient on its own to ensure sustained use of new technologies. Ongoing technical support, ideally integrated into the workflow, is needed to reduce barriers and facilitate continuous use.

Second, social and hierarchical dynamics within teams influenced implementation. Some therapists felt hesitant to introduce VR as a treatment option, fearing they lacked the authority to promote its use within their team. This suggests that, beyond technical and organizational barriers, workplace culture and professional autonomy play a crucial role in determining whether therapists feel empowered to implement new technologies. Studies have shown that the culture within a healthcare organization can either promote or hinder innovation. For instance, a study highlighted that cultural values can support innovation, but conflicts between existing culture and new ideas can restrain it. The manager’s role is crucial in nurturing this culture, indicating that leadership can significantly influence implementation outcomes [55].

Finally, a lack of organizational vision and sustained support following the training meant that VR was not naturally incorporated into routine treatment practice. Without clear structural reinforcement, such as time allocation, access to equipment, and leadership-driven implementation strategies, therapists were unlikely to prioritize VR over their usual treatment methods. These findings highlight the need for ongoing organizational commitment and support, addressing both technical and cultural barriers, to ensure the successful integration of VR into mental healthcare [56, 57]. It is important to note that the influence of leadership and organizational culture cannot be confined to one-off statements or actions. In this study, it became clear that support from management must be consistently demonstrated and integrated into the ongoing implementation process. It is not enough for leadership to endorse the technology at the outset; they must continue to champion its use, reinforcing that innovation is a priority. This ongoing support is essential to overcome resistance and to ensure that the implementation process is successful in the long term These findings indicate that training led to many positive changes in key implementation determinants. However, once the training was completed, new challenges emerged, highlighting the need for additional implementation strategies – particularly those focused on organizational support. A broader and more sustainable implementation process is required for successful VR implementation. Initially, the focus should be on training, but over time, other factors such as organizational support and leadership involvement become more critical. This shift emphasizes the importance of longitudinal evaluation and process monitoring, as challenges evolve. Furthermore, sustaining management’s engagement throughout the process is essential to maintain momentum and address emerging barriers, ensuring long-term success. This aligns with previous research demonstrating that behavior change in clinical practice requires more than training and must include elements of organizational vision, leadership support, and structural facilitation [56, 57].

### Strengths & Limitations

The findings of this study highlight the value of a longitudinal approach to studying implementation processes, as it captures the iterative and evolving nature of implementation efforts. By tracking changes over time, this approach provided unique insights into how key implementation determinants evolved and offered practical implications for adapting the implementation process in response to these changes. The longitudinal study design, therefore, plays a crucial role in exploring how experiences and context shift over time, underscoring its added value in understanding and guiding implementation processes [34, 58].

The iterative nature of the study also highlights the importance of formative evaluation; not only assessing implementation efforts at the end, but also continuously evaluating throughout the implementation process. This enables the identification of changing needs, ensuring that the implementation strategy can be adapted accordingly. Given the scope and the iterative nature of the interviews conducted across three time points, the sample size was sufficient to capture a comprehensive range of perspectives and experiences, thereby ensuring meaningful insights into the implementation process [59].

Alongside the iterative design, the use of the combined TDF and COM-B model contributed to a systematic and structured analysis of the implementation determinants. The model’s ability to distinguish between individual and contextual factors in behavior change proved particularly valuable in understanding the complexity of VR implementation [39].

However, the use of these models also comes with some limitations. The broad categorization within the capability, motivation, and opportunity categories could oversimplify the complex interactions between these determinants. Specifically, the opportunity category could oversimplify the complex interactions within an organization. Additionally, it can be difficult to clearly distinguish between some determinants in the data, such as between beliefs about capabilities and actual skills. These challenges highlight the need for further refinement of the models to capture the nuanced relationships and dynamic nature of implementation processes more accurately.

Alongside the limitations of the model, several other limitations should be noted. First, the perspective in this study is limited to therapists; managers, policymakers, trainers, patients, and other stakeholders were not interviewed. A broader perspective would likely have provided a more comprehensive understanding of the implementation determinants identified [60, 61]. However, focusing on therapists allowed for an in-depth exploration of their experiences, which is crucial for understanding practical implementation challenges as experienced by a main user group.

A related limitation is that there is a lack of generalizability. This study was conducted within one mental health organization, which may limit the direct applicability of the findings to other organizations of healthcare contexts. However, the insights gained regarding the role of time in the implementation process, such as how challenges evolve as the process unfolds and how implementation strategies should adapt to that, are likely to be relevant across different healthcare settings. The longitudinal nature of this study provides valuable implications for understanding how implementation dynamics change over time, which could inform similar initiatives in other organizations [62, 63]. Future research could adopt a multi-stakeholder approach and include cross-organizational comparisons of implementation strategies to further explore the nuances of VR implementation in diverse mental healthcare environments.

## Conclusion

The implementation strategy of VR training improves knowledge and skills but does not, on its own, lead to sustained use in clinical practice. Organizational and technical challenges pose significant barriers to implementation, with the lack of organizational vision and continuous practical support limiting progress. Therefore, VR training must be integrated into a broader and more sustainable implementation process. Successful implementation requires an iterative approach, where training, policy development, practical experiences, and formative evaluations work in tandem. The longitudinal design of this study added value by capturing how implementation determinants evolve, ensuring real-time needs from practice are continuously explored, and implementation strategies can be adjusted accordingly. Ultimately, stimulating the sustained implementation of VR in mental healthcare settings.

## Data Availability

The data underlying this study consist of anonymized interview transcripts. Due to ethical considerations and the sensitive nature of the content, these data cannot be shared publicly. However, anonymized transcripts can be made available upon reasonable request to the corresponding author, for research purposes only and subject to approval by the ethics committee of the University of Twente.

## Acknowledgments

We acknowledge the valuable contributions of all authors to this study. MK, HK, YB, and SK were involved in the study set-up, MK was involved in the data collection, MK and LK were involved in the data analysis, and MK was involved in the drafting of the manuscript. All authors have read and given feedback on the manuscript. All approved the final version of the manuscript. We would also like to thank Stefanie Gijsbertsen for her support throughout the research process. This study was funded by Stichting Vrienden van Oldenkotte (grant number: P2021-17 Transfore UT-FoReTech 2 Verder Vooruit). They had no role in the study design, data collection and analysis, decision to publish, or preparation of the manuscript.

## CRediT authorship contribution statement

Marileen M.T.E. Kouijzer: Writing – original draft, Conceptualization, Investigation, Methodology, Formal analysis, Visualization. Laura A.M. Koenis: Formal analysis, Writing – review & editing. David Huizinga: Conceptualization, Writing – review & editing. Saskia M. Kelders: Conceptualization, Supervision, Writing – review & editing. Yvonne H.A. Bouman: Conceptualization, Supervision, Writing – review & editing. Hanneke Kip: Conceptualization, Supervision, Writing – review C editing.

## Abbreviations

CFIR: Consolidated Framework for Implementation Research
COM-B: Capability, Opportunity, Motivation – Behavior model
EMDR: Eye Movement Desensitization and Reprocessing
NASSS: Non-adoption, Abandonment, and challenges to Scale-Up, Spread, and Sustainability
TDF: Theoretical Domains Framework
VR: Virtual Reality

**S1. Standards for Reporting Implementation Studies (StaRI) checklist**

